# Antimicrobial Resistance and Microbiological Gap Analysis for Central Nervous System Infections Bacterial Pathogens in Nigeria

**DOI:** 10.1101/2025.08.12.25333494

**Authors:** Jafar Eyad, Nouruldeen Saad, Amjad Benibella, Ramadhani Chambuso

## Abstract

**Background:** Despite extensive vaccine efforts, central nervous system (CNS) infections remain a significant cause of morbidity and mortality in the African meningitis belt. This is increasingly complicated by the rising antimicrobial resistance (AMR) in pathogens not covered by the vaccine. We mapped pathogen-specific AMR profiles and diagnostic gaps in CNS bacterial pathogens to inform precision microbiological-based interventions for improved surveillance and empiric therapy in Nigeria.

**Methods:** We conducted a retrospective analysis of a three year national AMR surveillance data, focusing on CNS bacterial pathogens. Data from 25 sentinel laboratories were extracted, analysed and interpreted per CLSI/GLASS standards. AMR profiles and diagnostic gaps were assessed using descriptive statistics and Chi-square tests for demographic and temporal comparisons.

**Results:** Among all 84,548 valid cultures from over 26,000 patients, culture positivity rate was higher in females (35.1%) and older adults (>65 years, 40.0%, **p < 0.001**). CSF specimens were underrepresented, while species-level ambiguity was high for CNS-associated infections. *Staphylococcus aureus*, *Escherichia coli* and *Klebsiella species* were the dominant isolates, while *Pseudomonas aeruginosa* and *Acinetobacter species* showed persistent low-level presence. Alarmingly, *Streptococcus pneumoniae* showed rising penicillin resistance, reaching 100% by 2018. Other pathogens, including *Klebsiella species*, *E. coli*, and *Pseudomonas aeruginosa* showed high AMR across multiple drug classes. Gap analysis scored all CNS associated bacterial pathogens at maximum clinical risk (5/5), with major deficits in detection and the laboratory capacity. A precision-targeted recommendation map tailored microbiological interventions, such as neonatal AST protocol for *E. coli* and ICU infection registries for *Pseudomonas species*.

**Conclusion:** Species-level identification gaps and high AMR in CNS infections bacterial pathogens demand targeted microbiological-led diagnostics to include expanded CSF testing and AST-guided empiric therapy in resource-limited settings.

**Highlights:**

- First national AMR gap analysis for CNS pathogens across 84,000 cultures in Nigeria
- Links resistance trends with diagnostic gaps for tailored stewardship interventions
- Introduces scalable microbiology-driven precision surveillance for LMIC settings
- Informs treatment protocols, lab policies and strategies for high-risk CNS infections

**Graphical abstract:** 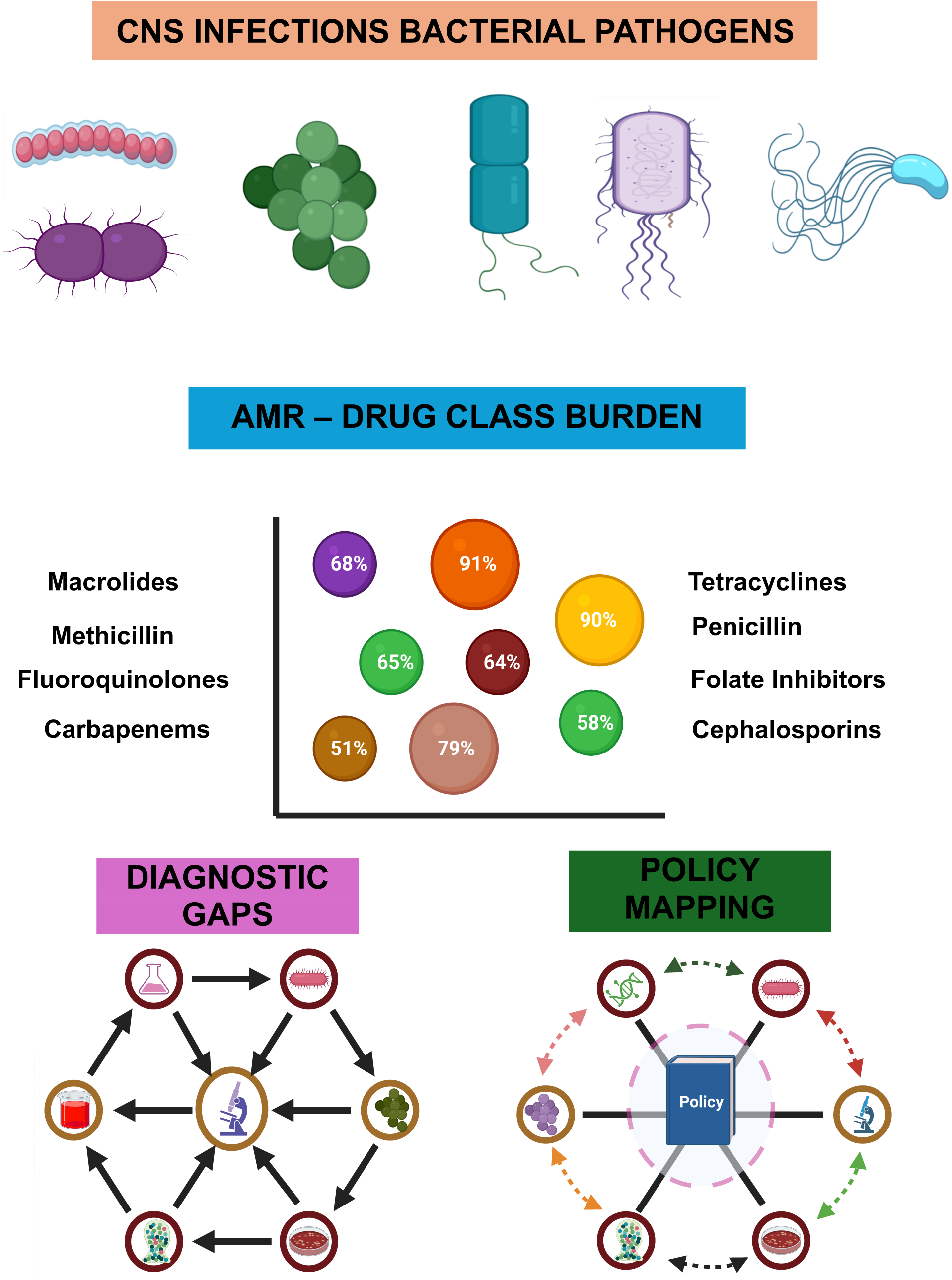

## Introduction

Central nervous system (CNS) infections are a group of serious and potentially life-threatening conditions that affect the brain, spinal cord, optic nerves, and their covering membranes.^1^ The CNS bacterial infections caused over 318,000 deaths globally each year, with sub-Saharan Africa accounting for more than half of this burden concentrated in the “African Meningitis Belt” which include countries like Nigeria, the most populous country within the “African Meningitis Belt”.^2,3^ For instance, between October 2022 and April 2023, Nigeria reported 1,686 suspected meningitis cases with 124 deaths, a case fatality rate of 7%.^4^ Existing vaccines such as the MenAfriVac have substantially reduced serogroup A *meningococcal* disease.^5,6^ However, other CNS bacterial infections remain driven by non-vaccine serogroups and resistant pathogens like *Streptococcus pneumoniae*, *E.coli*, and *Klebsiella pneumoniae*, where empirical treatment failure is increasingly linked to antimicrobial resistance (AMR).^7-10^

AMR is a parallel emergency in CNS bacterial infections, reducing the effectiveness of penicillins, cephalosporins, and fluoroquinolones, drugs essential for empirical therapy.^11^ Empirical therapy is further compromised by the absence of detailed national AMR microbiological mapping for CNS bacterial pathogens.^12,13^ Bacterial pathogens such as *Acinetobacter baumannii* and *Pseudomonas aeruginosa* are rarely reported in CNS infections, creating a false sense of security.^14,15^ Security is also lacking in species-level identification and laboratory capacity, leaving surveillance for CNS bacterial infections fragmented. Fragmentation has prevented a systematic gap analysis of CNS bacterial pathogens and AMR profiles.^16,17^ This resulted in a novel question on what are the pathogen-specific diagnostic, AMR and existing microbiological gaps undermining effective management of CNS bacterial infections, to address a critical and long-standing knowledge void.^18,19^

The identified knowledge void limits understanding of diagnostic blind spots with AMR profiles across high-associated bacterial pathogens for CNS infections.^20^ Without this knowledge, targeted interventions and empiric therapy will remain unfeasible and misaligned.^21^ Also, species-level detection is particularly important to address any rising resistance among neuroinvasive pathogens which further complicates the management of CNS infections.^22^ We hypothesize that CNS infections associated bacterial pathogens are disproportionately affected by detection capacity and improper AMR profiles which can be systematically mapped to inform microbiology-driven precision AMR strategies, tailored for national infection control programs.

To address our hypothesis in a global health perspective, we performed a microbiology-driven gap-analysis study of AMR in CNS infections for associated bacterial pathogens across a large-scale national surveillance data from Nigeria.

## Methodology

### Ethics statement

This study utilized publicly available, de-identified AMR surveillance data originally collected by the Nigerian Ministry of Health under the Fleming Fund Regional Grant (Phase 1) https://aslm.org/wp-content/uploads/2023/07/AMR_REPORT_NIGERIA.pdf?x89467. No individual-level data were accessed. Secondary data use complied with the Declaration of Helsinki and did not require additional ethical clearance.

### Study design

We conducted a retrospective study of the national AMR surveillance report of Nigeria for the years 2016 to 2018. This report, published in 2022, was generated through the Mapping Antimicrobial Resistance and Antimicrobial Use Partnership (MAAP) and involved 25 sentinel laboratories across Nigeria with bacteriology testing capacity. This study adheres to the STROBE (Strengthening the Reporting of Observational Studies in Epidemiology) guidelines.

### Data sources and surveillance framework

The original data were collected by Nigerian health authorities in collaboration with MAAP partners. Surveillance activities were conducted across a national network of medical laboratories selected for their bacteriology testing capacity. A total of 25 laboratories contributed antimicrobial susceptibility testing (AST) data using WHONET software. Trained field teams retrieved laboratory records from both paper-based and digital systems. Where feasible, these records were linked with hospital databases for clinical metadata, including age, sex, and specimen type. Specimens for culture were collected from cerebrospinal fluid (CSF), blood, urine and others like; Abscess/Discharge/Pus/Swab/Wound where these pathogens would be isolated from.

### Data extraction

From the national AMR report, the following data were extracted:

i. Total number of participated laboratories
ii. Collected specimens per year
iii. Source of collected specimens
iv. Specimen type
v. Valid cultures
vi. Positive and negative cultures.
vii. Species-level breakdown
viii. AST results for antibiotics tested
ix. Patient demographics (age and sex)

These variables were compiled into a structured dataset to allow for year-by-year resistance analysis

### CNS infections bacterial pathogens

While the surveillance report captured a broad range of pathogens, this study specifically focused on the most prevalent and clinically significant CNS infections-associated pathogens observed in the report. These organisms cause bacterial meningitis, neonatal sepsis, brain abscesses, ventriculitis, and other CNS-related conditions in both adult and paediatric populations.^10,23-28^ This includes:

a. Common CNS infections bacterial pathogens
  i. *Neisseria meningitidis*
  ii. *Haemophilus influenzae*
  iii. *Streptococcus species*: *Streptococcus pneumoniae and Streptococcus agalactiae*.
b. Rare CNS infections bacterial pathogens
  i. *Staphylococcus species*: *Staphylococcus aureus and Staphylococcus epidermidis*
  ii. *Pseudomonas specie*: *Pseudomonas aeruginosa*
  iii. *Klebsiella species*: *Klebsiella pneumoniae, Klebsiella oxytoca and Klebsiella aerogenes*
  iv. *Escherichia specie*: *Escherichia coli*
  v. *Acinetobacter species*: *Acinetobacter baumannii, Acinetobacter haemolyticus and Acinetobacter lwoffii*
  vi. Other *Streptococcus species* such as: *Streptococcus pyogenes, Streptococcus viridans, Streptococcus anginosus, Streptococcus bovis, Streptococcus gallolyticus, Streptococcus gordonii, Streptococcus milleri, Streptococcus mitis, Streptococcus oralis, Streptococcus parasanguinis, Streptococcus salivarius, Streptococcus sanguinis* and *Streptococcus suis*.

### Inclusion criteria and data management

Only isolates from the above list with valid AST results were included in the analysis. In accordance with WHO Global AMR Surveillance System (GLASS) guidelines and Clinical and Laboratory Standards Institute (CLSI) M39-A4 recommendations, only the first isolate per patient per year was retained to prevent duplicate entries.^29,30^ When unique patient identifiers were unavailable, all isolates were retained and interpreted with caution. AST interpretations followed CLSI guidelines, and WHONET interpretive rules were applied to ensure consistent reporting across laboratories.^31^

### AST and quality control

AST was performed using disk diffusion and MIC panels, depending on laboratory capacity, as described in the original report (https://aslm.org/wp-content/uploads/2023/07/AMR_REPORT_NIGERIA.pdf?x89467). CLSI standards were applied across all laboratories. Internal quality control was conducted using standard QC strains. External quality assessment (EQA) was implemented through structured proficiency testing coordinated by MAAP and the Fleming Fund. Although laboratories were encouraged to participate regularly, specific participation rates and EQA performance outcomes were not consistently detailed in the original report.

### AMR calculation

From the original report, AMR rates were derived from positive cultures with available AST results. AMR rates were calculated as the proportion of non-susceptible isolates (intermediate or resistant) relative to the total number of tested isolates within a single calendar year.

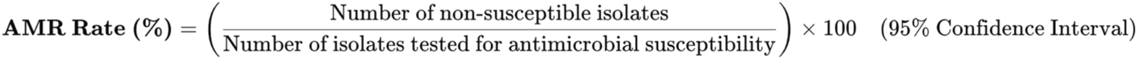

Where:

- *Non-susceptible isolates* = Resistant + Intermediate
- *Tested isolates* = All isolates subjected to AST

### Statistical analysis

Analyses were carried out in R version 4.3.2. Because less than 1% of the data was missing, a complete-case analysis was deemed sufficient. Valid culture data were analyzed using descriptive statistics to explore positivity rates by demographic group and by year and Wald confidence intervals (CI) were calculated. Pearson’s Chi-square test was used for categorical comparisons, with p < 0.05 considered statistically significant.

## Results

### 1. Demographics and culture characteristics

Among the 84,548 valid cultures processed across all laboratories for all specimens, culture positivity was notably higher in females (35.1%) than males (28.3%), with a significant statistical difference across gender groups (***p* < 0.001**). Older age groups showed increased positivity, rising from 23.9% in children aged 1–17 years to 40.0% in those above 65 years, with the highest yield seen in the unknown age group (42.5%; ***p* < 0.001**). Over the years, culture positivity steadily increased from 28.5% in 2016 to 34.6% in 2018 (***p* < 0.001**), suggesting improved diagnostic recovery or changing epidemiology.

**Table 1.** Diagnostic yield across all cultures.

| Variable | Group | Valid cultures, n | Positive cultures with AST, n | Negative cultures, n (%) | Positive cultures, n (%) | Negative cultures, n (%) | 95% CI for Positive (%) | *p-value |
| --- | --- | --- | --- | --- | --- | --- | --- | --- |
| Gender | Male | 37550 | 10160 | 26932 (71.7%) | 10618 (28.3%) | 26932 (71.7%) | (27.8 - 28.7) | <0.001 |
|  | Female | 46997 | 13803 | 30480 (64.9%) | 16517 (35.1%) | 30480 (64.9%) | (34.7 - 35.6) |  |
| Age | < 1 year | 9689 | 2711 | 6857 (70.8%) | 2832 (29.2%) | 6857 (70.8%) | (28.3 - 30.1) | <0.001 |
|  | 1 – 17 yrs | 20476 | 4526 | 15714 (76.7%) | 4902 (23.9%) | 15714 (76.7%) | (23.4 - 24.5) |  |
|  | 18 – 49 yrs | 29336 | 7629 | 20013 (68.2%) | 9323 (31.8%) | 20013 (68.2%) | (31.2 - 32.3) |  |
|  | 50 – 65 yrs | 5177 | 1630 | 3424 (66.1%) | 1753 (33.9%) | 3424 (66.1%) | (32.6 - 35.2) |  |
|  | > 65 yrs | 4743 | 1806 | 2845 (60.0%) | 1898 (40.0%) | 2845 (60.0%) | (38.6 - 41.4) |  |
|  | Unknown | 15127 | 5661 | 8700 (57.5%) | 6427 (42.5%) | 8700 (57.5%) | (41.7 - 43.3) |  |
| Year | 2016 | 26896 | 6532 | 19218 (71.5%) | 7678 (28.5%) | 19218 (71.5%) | (28.0 - 29.1) | <0.001 |
|  | 2017 | 32134 | 9501 | 21494 (66.9%) | 10640 (33.1%) | 21494 (66.9%) | (32.6 - 33.6) |  |
|  | 2018 | 25518 | 7930 | 16701 (65.4%) | 8817 (34.6%) | 16701 (65.4%) | (34.0 - 35.1) |  |

\*Pearson's $\chi^2$ test comparing the distribution of positive versus negative cultures across all categories within each variable.

### 2. Diagnostic yield, specimen distribution and temporal trends of all cultures across participated laboratories

There were critical diagnostic gaps in CNS specimen collection and species resolution, particularly for high-associated neurological pathogens (**Fig 1**). We observed inter-laboratory variability in diagnostic and AST coverage was substantial, with some high-volume centers exceeding 90% AST reporting while others, such as Bwari and FNHI Yaba, showed critical shortfalls (**Fig 1 a**). Among specimen types, CSF accounted for only 209 samples, making it the least submitted despite its importance for CNS diagnostics (**Fig 1b and d**). *Cryptococcus neoformans*, *Neisseria meningitidis* and *Listeria monocytogenes*, key CNS pathogens were rarely isolated (**Fig 1 c**). Species-level ambiguity was high for *Neisseria* (95%) and *Cryptococcus* (90%), severely limiting diagnostic confidence in CNS infections (**Fig 1h**). *Staphylococcus aureus* remained the leading isolate overall, peaking in 2017, while *Escherichia coli* and *Klebsiella pneumoniae* showed consistent but variable prevalence (**Fig 1 c, e and g**). Although *Pseudomonas aeruginosa* and *Acinetobacter baumannii* were less frequent, their stable presence signals persistent risk of nosocomial CNS infections.^32^

**Fig 1.**
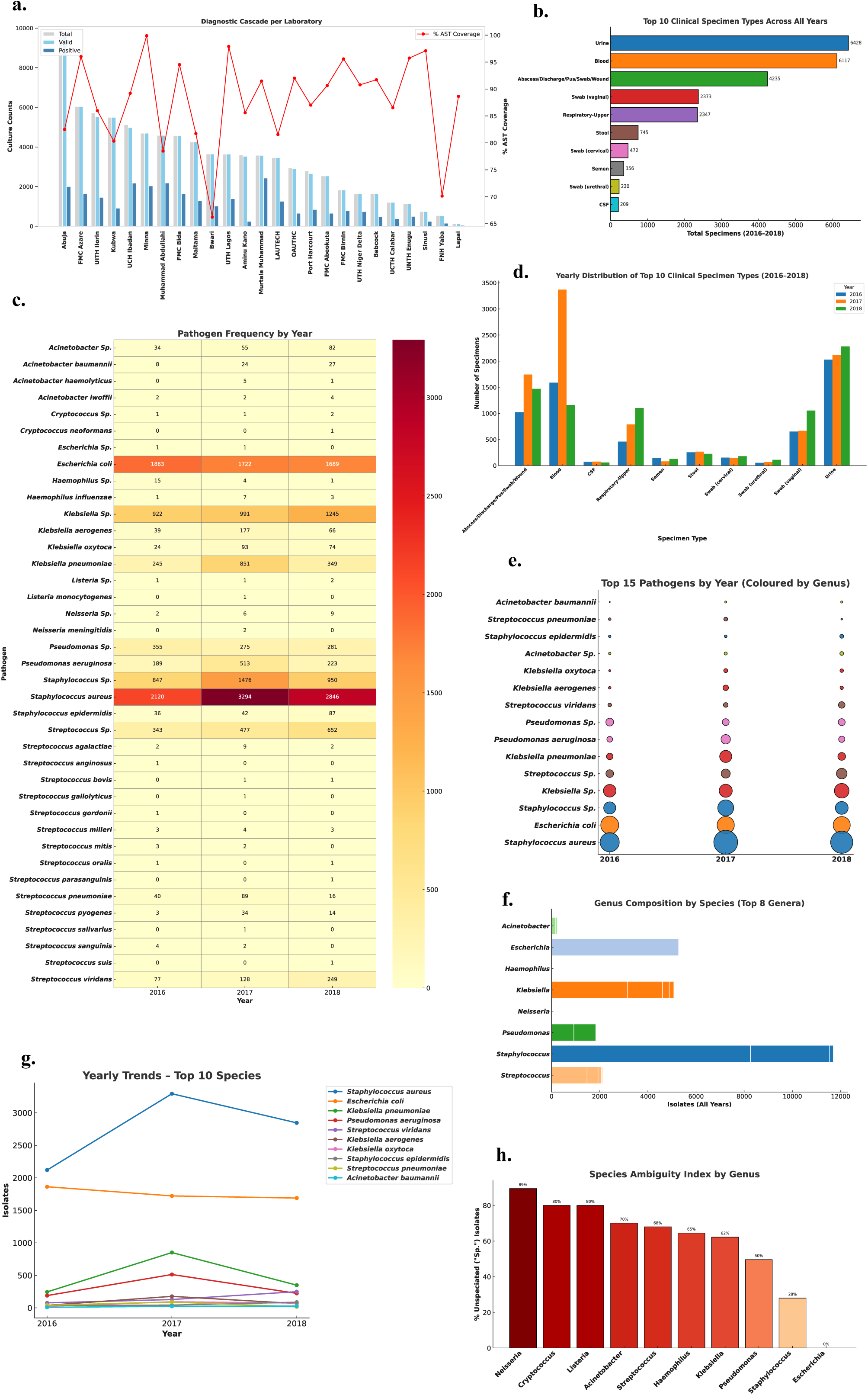
Diagnostic and epidemiological landscape across multiple laboratories and specimen types in Nigeria. (**a**) Diagnostic cascade per laboratory showing counts of total, valid and positive cultures, overlaid with % AST coverage (red line), highlighting inter-laboratory differences in diagnostic performance and antimicrobial susceptibility testing. (**b**) Urine, blood, and abscess/pus/wound specimens were the most frequently submitted specimen types across all years. (**c**) Heatmap of pathogen frequency by year indicating *Staphylococcus aureus* and *Escherichia coli* as dominant pathogens, with temporal fluctuations in *Klebsiella pneumoniae*, *Pseudomonas aeruginosa*. (**d**) Yearly distribution of the top 10 clinical specimen types showing increased blood and urine specimen submissions over time and underrepresentation of CSF for CNS infections. (**e**) Top 15 pathogens stratified by year and genus showing *S. aureus*, *E. coli* and *Klebsiella* spp. as consistently prevalent species. (**f**) Genus-level composition of isolates illustrating *Staphylococcus* dominance and species diversity within *Klebsiella* and *Escherichia*. (**g**) Yearly trends for the top 10 species showing peaks in *S. aureus* and *K. pneumoniae* during 2017. (**h**) Species ambiguity index by genus showing high rates of non-speciated (“sp.”) entries among *Neisseria*, *Cryptococcus*, *Listeria*, and *Acinetobacter*, emphasizing limitations in species-level identification for key CNS pathogens.

### 3. Yearly AMR patterns across drug classes and specific CNS pathogen species

The AMR among key CNS infections bacterial pathogens revealed escalating threats to empirical treatment, particularly for *Streptococcus pneumoniae*, a major cause of meningitis, exhibited alarmingly high to complete resistance to penicillin, rising from 67% in 2016 to 100% in 2018, alongside persistently elevated macrolide resistance (>70%), undermining first-line CNS therapy (**Fig 2**). *Staphylococcus aureus* and *E.coli* show high resistance of folate pathway inhibitors. Among Gram-negative pathogens, *Klebsiella* species displayed high resistance to 3^rd^ generation cephalosporins. *Pseudomonas species* showed high resistance to tetracyclines (86%).

**Fig 2.**
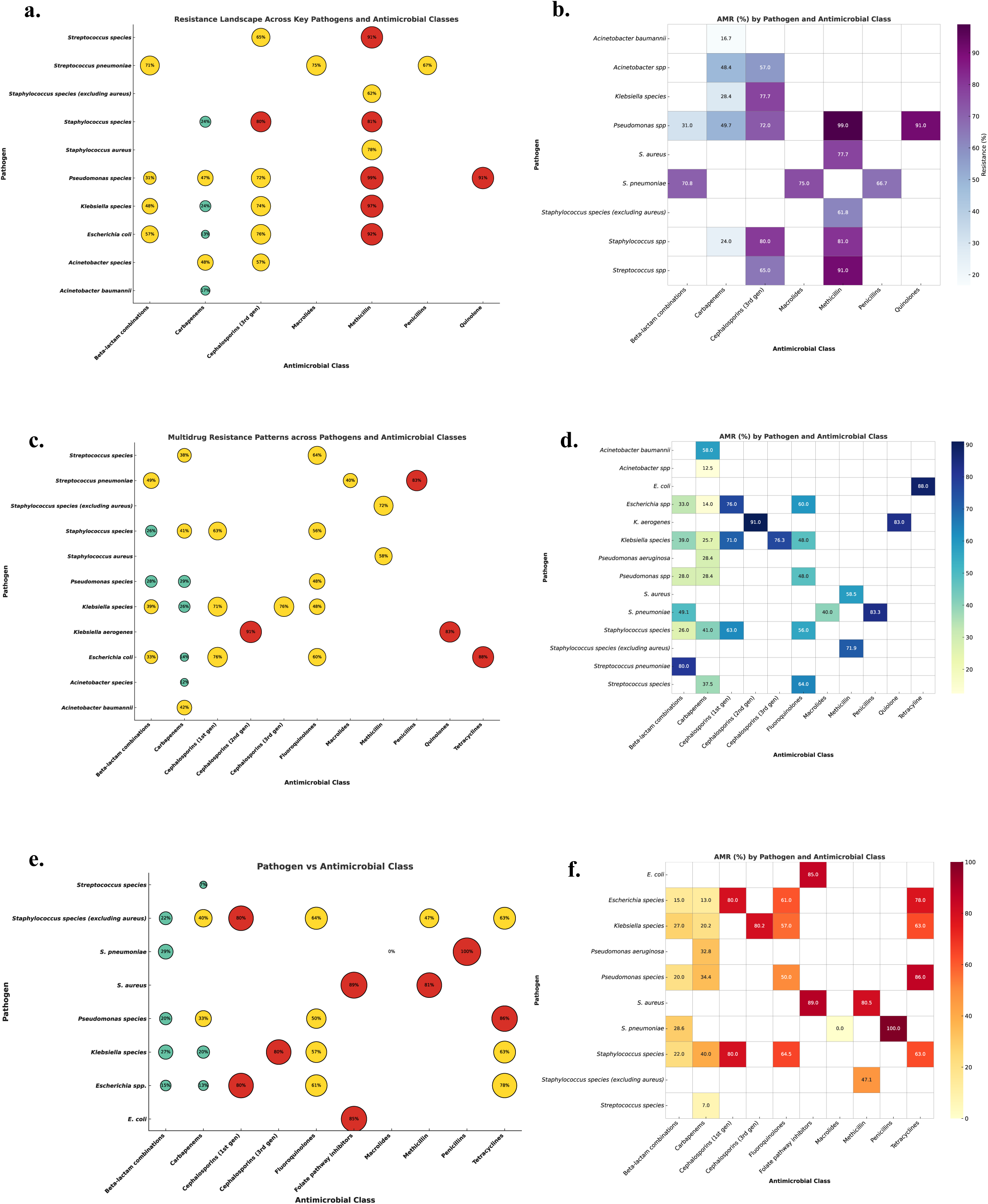
AMR profiles of associated CNS pathogens across different drug classes. **(a)** Bubble plot for year 2016 showing widespread high resistance for 3^rd^ generation cephalosporins, and methicillin, while high fluoroquinolone resistance in *Pseudomonas species* (91%). (**b**) Heatmap showing similar patterns. **(c)** Bubble plot for 2017 highlights emerging resistance in *Klebsiella aerogenes* (91% to 2^nd^ generation cephalosporins, and quinolones 83%) and *E. coli* (88% to tetracyclines), with *Streptococcus pneumoniae* showing 83% resistance to penicillin. (**d**) 2017 heatmap showing high resistance in *Klebsiella aerogenes* to 2^nd^ generation cephalosporins (91%). (**e**) Bubble plot for 2018 indicates *S. pneumoniae* had complete (100%) penicillin resistance. (f)A 2018 heatmap illustrating similar trend signalling critical gaps in effective empiric therapy for CNS and systemic infections.

### 4. AMR analysis for CNS pathogens to address critical microbiological-related gaps

Gap analysis across eight CNS-relevant associated pathogens revealed critical deficiencies in detection, resistance profiling, and laboratory capacity, with consistently high clinical risk scores (5/5) across all organisms (**Fig 3**). *Streptococcus pneumoniae* and *Staphylococcus aureus* scored maximally for resistance and risk, reflecting the burden of penicillin-resistant meningitis and invasive MRSA infections, respectively, both compounded by low lab capacity. CNS device-associated pathogens (*Staph/Strep species*) and *Acinetobacter baumannii* also demonstrated high resistance and inadequate detection infrastructure, highlighting vulnerabilities in neurosurgical infection control in Nigeria.^32^ Surveillance for CNS pathogens overall scored worst in detection gap (5/5), exposing systemic underreporting and incomplete data across sentinel laboratories. *Klebsiella species* and *Escherichia coli*, both linked to severe neonatal CNS infections showed extreme resistance (5/5). Both *Pseudomonas aeruginosa* and *Acinetobacter* species showed elevated resistance and CNS risk, further reinforce the urgent need for targeted diagnostics and empiric therapy realignment in CNS infections.

**Fig 3.**
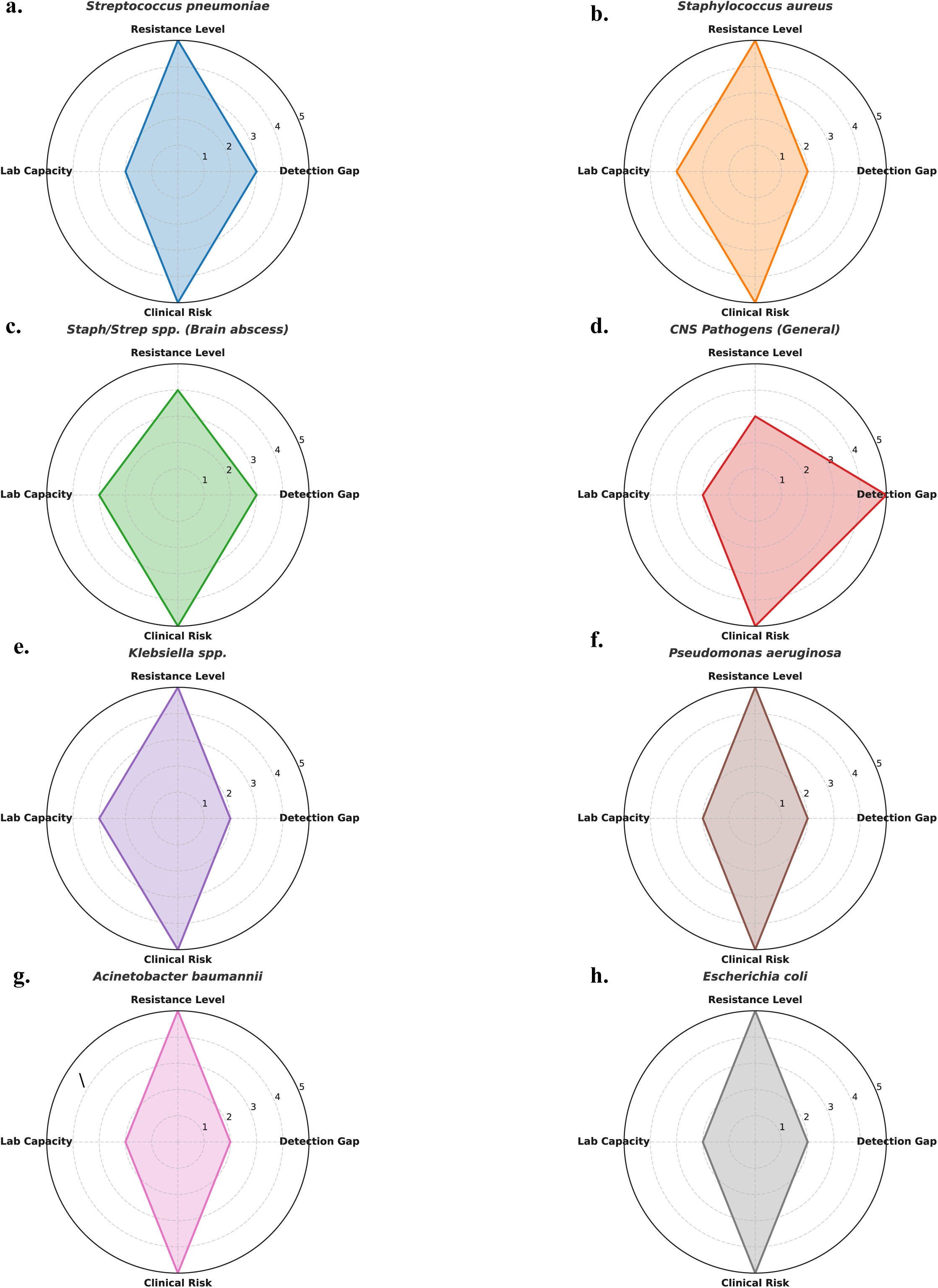
Pathogen-specific gap analysis for CNS-relevant high-associated organisms across four domains: detection gap, resistance level, laboratory capacity, and clinical risk. (**a**) *Streptococcus pneumoniae* shows maximal resistance and clinical risk scores (5/5), with poor laboratory capacity (2/5) and a moderate detection gap (3/5). (**b**) *Staphylococcus aureus* shows moderately lab capacity in screening and low detection gap. (**c**) *Staph/Strep spp.* associated with brain abscess exhibited high clinical risk and resistance level but are affected by moderate detection and lab capacity. (**d**) CNS pathogen surveillance scores worst in detection (5/5) and lab capacity (2/5). (**e**) *Klebsiella species* demonstrate high resistance (5/5) and clinical risk and low detection gap. (**f**) *Pseudomonas aeruginosa* exhibits uniformly high resistance and clinical risk with low detection and lab gaps (2/5 each). (**g**) *Acinetobacter baumannii* show equally elevated resistance and clinical risk scores with low detection and lab capacity. (**h**) *Escherichia coli* displays high resistance levels and clinical risk while showing lab and detection limitations.

### 5. Addressing identified pathogen-specific challenges for CNS infections

We explored a precision surveillance strategy, visually aligning pathogen biology with targeted diagnostics, stewardship, and infection prevention across diverse CNS clinical settings (**Fig 4**). This was performed to provide a strategic, organism-specific action model to inform diagnostic stewardship and infection prevention frameworks for CNS infections in high-burden, resource-limited settings. Targeted intervention mapping across eight high-associated CNS pathogens revealed key microbiology-informed gaps in AMR response systems. *Escherichia coli*, frequently implicated in neonatal meningitis, was recommended to initiate neonatal AST protocol while *Klebsiella species* and *Pseudomonas aeruginosa*, both frequent in post-surgical CNS infections, were recommended for quarterly AMR feedback and ICU-based infection registries, respectively. *Acinetobacter baumannii*, a notoriously persistent MDR pathogen in neurocritical care, was recommended to initiate a CNS registry.^32^ Gram-positive cocci including *Staphylococcus aureus* and *Streptococcus pneumoniae* were distinctly tied to IPC bundles and AST lab geo-planning, reflecting their dual role as colonizers and invasive agents. Notably, CNS-wide diagnostic gaps were addressed through the inclusion of sentinel AMR laboratories, emphasizing a systems-level response to data fragmentation.

**Fig 4.**
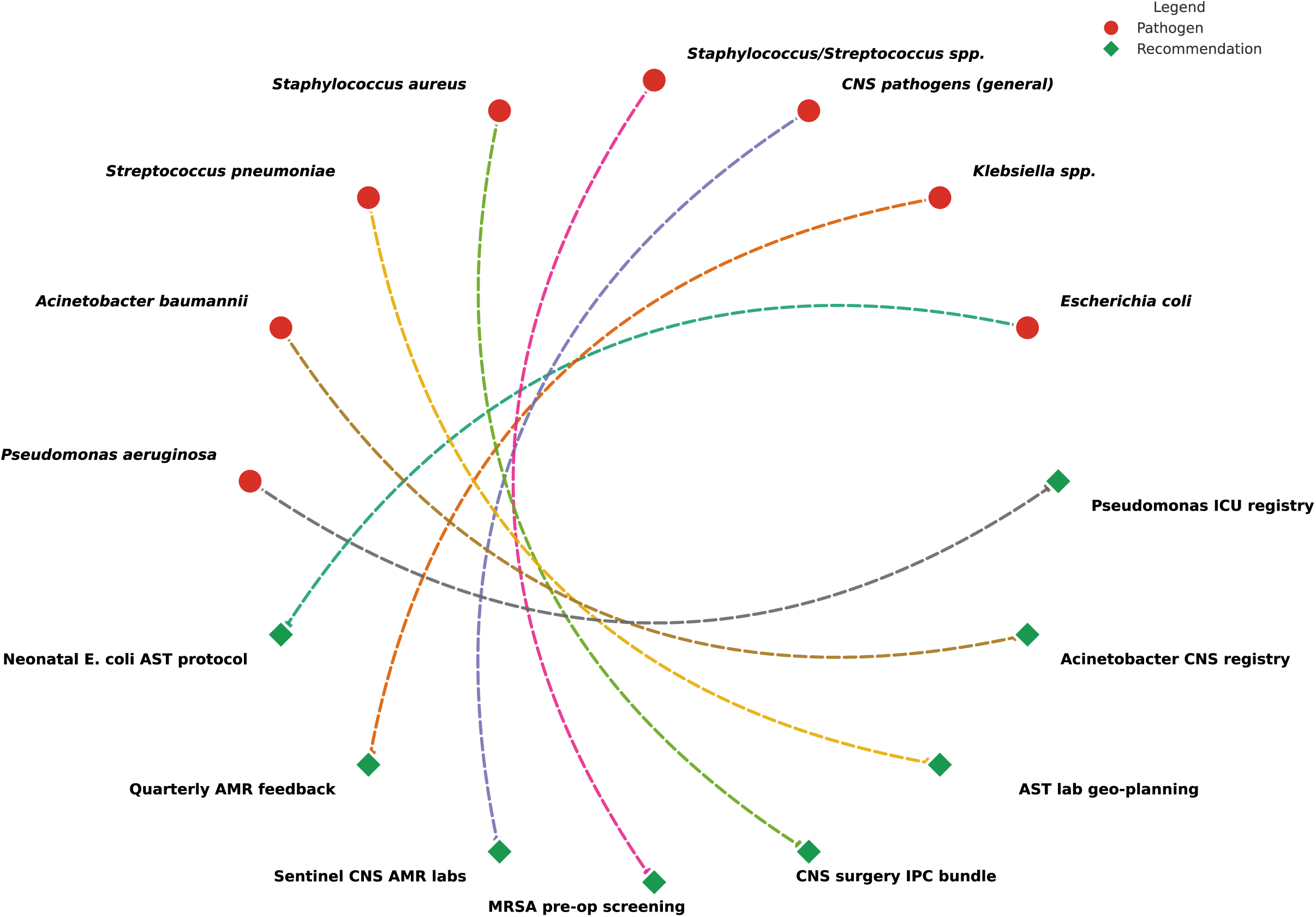
Pathogen-specific recommendations for AMR strategies for CNS infections. A circular network maps eight associated CNS pathogens to tailored microbiological interventions. Recommendations include *Escherichia coli* with neonatal AST protocols, *Acinetobacter baumannii* with CNS registry and *Pseudomonas aeruginosa* with ICU-based CNS infection registries. Cross-cutting gaps were addressed by linking general CNS pathogens to sentinel AMR laboratories with molecular diagnostics.

## Discussion

This study provides the first integrated microbiological-driven gap-analysis assessment of AMR in CNS infections associated pathogens across a large-scale national surveillance platform in Nigeria, encompassing over 84,000 cultures. By linking organism-specific resistance trends, diagnostic performance gaps and pathogen-tailored recommendations, our study introduces a precision AMR surveillance model uniquely adapted for resource-limited settings. The novel use of structured gap analysis and a creative intervention mapping for CNS infections associated pathogens including *Escherichia coli*, *Klebsiella species*, *Pseudomonas aeruginosa*, *Staphylococcus aureus* and *Streptococcus pneumoniae* translates complex laboratory surveillance data into actionable practical tools such as neonatal AST protocols, ICU infection registries and IPC bundles. These findings offer a scalable framework to inform empiric CNS infections treatment guidelines, optimize lab resource allocation, and strengthen national AMR policy. Given the global urgency of rising AMR in CNS pathogens and the absence of structured diagnostic and stewardship strategies in many LMICs, this study fills a critical evidence gap and has direct implications for regional and global CNS infections preparedness.

The current study emphasises on the profound microbiological vulnerabilities in the surveillance and management of CNS infections, particularly in how diagnostic insufficiencies amplify the impact of AMR in Nigeria.^33,34^ The dominance of non-speciated pathogen genera reflects not only a failure of species-level diagnostics but also a missed opportunity to implement pathogen-specific treatment and public health interventions, especially for non-vaccine-preventable infections. High and rising resistance rates in *Streptococcus pneumoniae* and *Klebsiella species* suggest selective pressure from empiric therapy regimens that may be poorly matched to local susceptibility profiles, highlighting the urgency for local antibiogram-driven prescribing.^35^ In addition, organisms traditionally considered secondary in CNS infections such as *Acinetobacter baumannii* and *Pseudomonas aeruginosa* maintained a stable foothold, likely due to their resilience in healthcare environments and association with device-related infections.^32,36^ Moreover, the high prevalence of resistance to critical antimicrobial drug classes such as cephalosporins, tetracyclines and folate inhibitors across pathogens associated to CNS infections reflects the existing challenges facing antimicrobial armamentarium in both Gram-positive and Gram-negative CNS associated pathogens.^37,38^ In our study, we have exposed this significant microbiological landscape characterized by incomplete species resolution, over-reliance on empiric broad-spectrum therapy and inadequate alignment between laboratory evidence and clinical decision-making, challenges that are central to both treatment failure and the acceleration of resistance, globally.

The results of this study align with and extend prior regional AMR surveillance efforts while revealing critical divergences that expose the limitations of previously available data. For instance, the WHO GLASS report (2022) acknowledges rising resistance in *Streptococcus pneumoniae* and *Klebsiella pneumoniae* globally, but our data show that *S. pneumoniae* resistance to penicillin in CNS infections reached 100% by 2018, significantly higher than the 45–60% reported in aggregated GLASS datasets for sub-Saharan Africa.^39^ This discrepancy likely reflects the inclusion of rare CNS associated isolates in global estimates and highlights the unique resistance pressure within CNS compartments, where blood–brain barrier pharmacokinetics and delayed diagnosis may drive more aggressive antibiotic use.^40-42^ Similarly, our finding of 86% tetracycline resistance in 2018 for *Pseudomonas species* is considerably higher than the 53% resistance rate previously reported in Nigeria, likely reflecting nosocomial CNS-specific strains associated with device colonization and prolonged ICU stays.^32,33,43,44^ The diagnostic gaps observed here, especially the 95% species-level ambiguity contrast sharply with surveillance systems in high-income countries like the CDC’s Active Bacterial Core Surveillance (ABCs), which report >95% speciation accuracy due to universal adoption of PCR-based diagnostics and centralized data harmonization. Moreover, while most African AMR studies focus on urinary or bloodstream isolates, our CNS-specific gap analysis study uniquely exposes the underrepresentation of CSF samples and the lack of tailored response strategies for neurological infections. These differences emphasize the contextual nature of AMR dynamics and the need for surveillance systems that capture niche infection types and high-risk anatomical sites. The convergence of high resistance, diagnostic fragmentation and clinical risk in our findings thus presents a distinct epidemiological profile that current regional and global frameworks fail to adequately capture.^45^

The pathogen-specific gap analysis conducted in this study revealed not only the extent of diagnostic and resistance challenges but also offered a structured framework for microbiologically informed intervention. By quantifying detection gaps, laboratory capacity limitations, and clinical risk across eight CNS associated pathogens, the analysis exposed consistent deficits that disproportionately affect organisms with high virulence and poor therapeutic coverage. For instance, *Streptococcus pneumoniae* and *Staphylococcus aureus* exhibited maximal resistance and risk scores, yet were matched with suboptimal laboratory capacity, pointing to an urgent need for targeted AST strengthening and IPC programs. The translational strength of this study lies in its alignment of these quantitative gaps with actionable microbiological solutions, such as neonatal-specific *E. coli* AST protocols, *Acinetobacter* CNS surveillance registries and geospatial AST lab expansion for high-burden regions.^32^ Unlike generic AMR strategies, these recommendations are pathogen-matched and informed by real surveillance data, offering governments and hospital networks a prioritization model to guide resource allocation and diagnostic scale-up.^46,47^ Our approach transforms abstract surveillance outputs into a practical toolkit for strengthening empiric therapy frameworks, improving early detection of high-risk CNS infections, and institutionalizing precision AMR responses in neurosurgical and neonatal care settings.

The strengths that distinguish this study include:

i. First large-scale CNS-focused AMR surveillance study using secondary data which incorporates more than 84,000 cultures results with pathogen-specific AMR across multiple years and laboratories.
ii. Integration of laboratory analysis of diagnostic yield, species resolution and AST coverage exposes systemic weaknesses often missed in pathogen-only studies.
iii. Use of structured, pathogen-specific gap analysis study which quantifies detection, resistance, lab capacity and clinical risk across eight CNS associated pathogens in Nigeria, introducing a novel scoring framework for prioritizing interventions.
iv. Introduction of precision intervention mapping and first study to apply visualization to align CNS associated pathogens with tailored, microbiology-informed recommendations.
v. Our CNS infections-specific AMR patterns linked to empirical treatment implications which show high-level resistance in *S. pneumoniae*, *Klebsiella* and *Pseudomonas* directly challenges the current first-line CNS infection therapies in LMICs.
vi. The focus on underrepresented CNS associated pathogens from high-risk clinical samples (CSF), highlights the diagnostic neglect of CSF and CNS infections in national surveillance systems, guiding to re-consider clinical sample collection and lab policy for CNS infections.
vii. Provides data-driven tools to guide policymakers in laboratory investment, empiric guideline development and targeted infection control interventions for CNS infections.
viii. Provides a scalable framework adaptable to other LMICs seeking to build CNS pathogen surveillance within fragmented or underfunded clinical and laboratory networks.

The limitations of this study include:

i. Underrepresentation of CSF specimens since CNS infections were poorly captured due to the extremely low volume of CSF submissions, limiting pathogen diversity and clinical correlation.
ii. High species-level ambiguity for multiple CNS pathogens, undermining diagnostic specificity and epidemiological validity.
iii. Inter-laboratory heterogeneity in AST coverage that marked differences in AST reporting rates across laboratories which introduced potential biases in resistance estimates and reduced comparability.
iv. Absence or the lack of clinical outcome data (e.g., mortality, treatment response) restricts the ability to evaluate the direct impact of AMR patterns on patient care and treatment outcomes.
v. Cross-sectional surveillance retrospective design and the use of aggregated yearly data limits the ability to assess temporal causality or emergence of specific resistance clones over time.
vi. Limited molecular diagnostic data and absence of genotypic or PCR-based confirmation precludes detection of resistance genes, virulence factors, or outbreak clusters.

Despite these limitations, this study offers critical and actionable insights by leveraging one of the largest CNS-focused culture datasets in the region, revealing resistance patterns and diagnostic gaps that have not been previously characterized. The use of robust microbiological analysis, cross-laboratory comparison, and pathogen-specific intervention mapping ensures that the findings remain valid and highly relevant for national AMR response planning. Moreover, the identified gaps themselves emphasize the urgent need for surveillance reform, making the results not only valuable but essential for guiding future diagnostic investments and empirical treatment policies in low-resource settings.

## Conclusions

This study provides the first secondary data use for national microbiology-centered analysis of CNS-infection associated pathogens surveillance and AMR in Nigeria. It builds upon the foundational progress made by Nigerian public health authorities and surveillance laboratories by providing the first focused analysis of CNS infections through a microbiological and global health systems lens. It suggests that, despite substantial national investment in AMR surveillance in Nigeria, critical gaps persist in the detection, species-level identification and antimicrobial susceptibility testing of high-associated CNS pathogens. The findings highlight the urgent need to complement ongoing efforts with pathogen-specific, site-adapted strategies that strengthen laboratory capacity and empiric treatment guidance. By introducing a precision gap analysis and intervention model, our study offers a translational framework to enhance existing surveillance programs and inform national policy, without undermining the valuable progress already achieved. The key takeaway message is that national AMR response strategies must move beyond generic stewardship to pathogen-targeted, site-specific interventions. Immediate priorities should include expanding CSF diagnostics, adopting routine species-level identification and deploying AST-guided empiric therapy models, particularly for high-risk neurological and neonatal infections. These findings are scalable and urgently relevant to similar high-burden, resource-constrained settings across sub-Saharan Africa.

## Supporting information

Supplementary Data Set

## Data Availability

This study utilized publicly available, de-identified AMR surveillance data originally collected by the Nigerian Ministry of Health under the Fleming Fund Regional Grant (Phase 1) No individual-level data were accessed. Secondary data use complied with the Declaration of Helsinki and did not require additional ethical clearance.

https://aslm.org/wp-content/uploads/2023/07/AMR_REPORT_NIGERIA.pdf?x89467.

